# The evaluation of factors affecting antibody response after administration of the BNT162b2 vaccine: A prospective study in Japan

**DOI:** 10.1101/2021.06.20.21259177

**Authors:** Toshiya Mitsunaga, Yuhei Ohtaki, Yutaka Seki, Masakata Yoshioka, Hiroshi Mori, Midori Suzuka, Syunsuke Mashiko, Satoshi Takeda, Kunihiro Mashiko

## Abstract

The aim of this study was to evaluate the antibody reaction after administration of the BNT162b2 vaccine, and to reveal the factors that affect antibody production. This prospective study was carried out in the Association of EISEIKAI Medical and Healthcare Corporation Minamitama Hospital, in Tokyo, Japan, from April 15, 2021 to June 09, 2021. All our hospital’s workers who were administered the BNT162b2 vaccine as part of a routine program were included in this study.

We calculated the anti-SARS-CoV-2 spike-specific antibody titter 1) before vaccination, 2) seven to twenty days after the first vaccination, and 3) seven to twenty days after the second vaccination.

The low-antibody titer group (LABG) was defined as the group having less than 25 percentiles of antibody titer. Univariate and Multivariate logistic regression analysis were performed to ascertain the effects of factors on the likelihood of LABG. 374 participants were eventually included in our study, and they were divided into 94 LABG and 280 non-LABG. All samples showed significant antibody elevation in the second antibody test, with a mean value of 3476 U/mL. When comparing the LABG and non-LABG groups, the median age, blood sugar, and HbA1c were significantly higher in the LABG group. The rates of participants with low BMI (<18.5) and high BMI (>30) were significantly higher in the LABG group. The proportion of chronic lung disease, hypertension, diabetes, dyslipidemia, autoimmune disease, and cancer were significantly higher in the LABG group. Although there was no significant difference confirmed with respect to the exercise hours per day, the proportion of participants that did not perform outdoor activities was significantly higher in the LABG group. The time interval between the second vaccination and the second antibody test, and between the first and the second vaccination was significantly longer in the non-LABG group.

Our logistic regression analysis revealed that the age, obesity, hypertension, diabetes, dyslipidemia, antihypertensive drug, antilipid drug, γ-GT, BS, HbA1c, and lack of outdoor activity were significant suppressors of antibody reaction, whereas maintaining the appropriate time interval between the first and the second vaccination could promote a significant antibody response. In the multivariate logistic regression analysis, age, obesity, and lack of outdoor activities were significant suppressors of antibody reaction, whereas the length of days from the first to the second vaccination promoted a significant antibody response.

Our single-center study demonstrates that age, obesity, and lack of outdoor activities were significant suppressors of antibody response, whereas maintaining the appropriate time interval between the first and the second vaccination could promote a significant antibody response. Evidence from multi-center studies is needed to develop further vaccination strategies.

## Introduction

The severe acute respiratory syndrome coronavirus 2 (SARS-CoV-2), which started in Wuhan, China, has spread all over the world significantly faster than we expected. By the end of April 2021, the number of cases infected with SARS-CoV-2 had exceeded 150 million, with more than 3.1 million deaths (mortality rate: 2.1%) (***World Health Organization, 2021***). According to the the Ministry of Health, Labour and Welfare of Japan, the number of coronavirus 2019 (Covid-19) cases confirmed by 30 April 2021 in Japan was 588,900, with 10,226 deaths (mortality rate: 1.74%) (***The Ministry of Health, Labour and Welfare, 2021***.). Many developed countries in cooperation with the World Health Organization (WHO) have been trying to reduce the number of Covid-19 cases. However, because of its strong infectivity, it has been very challenging to control this disease.

The majority of patients infected with Covid-19 were mild cases, but approximately 5% of cases progressed to severe conditions, and approximately 2% died (***Li et al., 2020***.). Several studies have suggested that the risk factors for severe Covid-19 involve individual’s age, gender (males are at higher risk compared to females), smoking, and pre-existing conditions such as obesity, diabetes, chronic lung disease, hypertension, dyslipidemia, and chronic kidney disease (***Matsunaga et al., 2020; Mi et al., 2020; Liang et al., 2020; Lippi et al., 2020; Myers et al., 2020; Fadini et al., 2020; Zheng et al., 2020; Popkin et al., 2020***). Although several treatment strategies, including steroids and antiviral drugs, have been developed, vaccination is still the most important means of preventing Covid-19 infection and aggravation in people with these risk factors. BNT162b2 is a new generation vaccine with nucleoside-modified RNA molecules encoding the full-length SARS-CoV-2 spike glycoprotein (***Polack et al., 2020***). A previous study showed that the efficacy of this vaccine in preventing Covid-19 is approximately 95%, and thus extremely high (***Polack et al., 2020***). Typically, people have to be vaccinated twice to boost their immunity, and the study carried by Walsh *et al* showed that spike binding IgG titer increased dramatically and reached a plateau after only seven days from the second dose (***Walsh et al., 2020***). However, the major studies of Covid-19 vaccines are carried out in non-Asian countries, and the true efficacy of this vaccine in Asian people is still unclear (***Polack et al., 2020; Walsh et al., 2020***).

Certain studies have been identifying the factors that promote or suppress antibody response after vaccination for several viruses. A study performed by Pellini *et al* showed that obesity and age, which were the risk factors for severe Covid-19, may also be interfering factors for SARS-CoV-2 vaccine immunogenicity (***Pellini et al., 2021***). However, the population of this study was relatively small and only included non-Asian populations. In contrast, the studies performed by Tanja *et al* and Prather *et al* demonstrated that sleep duration and sleep quality, which reflect the circadian rhythm, were positively correlated to antibody production after Influenza vaccination or Hepatitis A vaccination, respectively (***Tanja et al., 2003; Prather et al., 2021***). However, to our knowledge, there have been no studies that evaluate the relationship between the factors involved in the daily life rhythm and the antibody response after Covid-19 vaccination.

Therefore, the aim of this study was to evaluate the anti-SARS-CoV-2 spike-specific antibody reaction following the administration of BNT162b2 vaccine, and to reveal the factors that affect antibody response in a Japanese population.

## Materials & Methods

### Study design

This prospective study was carried out between April 15, 2021 and June 9, 2021 at the Association of EISEIKAI Medical and Healthcare Corporation Minamitama Hospital, a secondary emergency medical institution. The protocol for this research project was approved by a suitably constituted Ethics Committee of the institution and conforms to the provision of the Declaration of Helsinki (Committee of Association of EISEIKAI Medical and Healthcare Corporation Minamitama Hospital, Approval No. 2020-Ack-19), and written consent was obtained from all the human subjects.

### Study setting and population

All our hospital’s workers, who were administered with the BNT162b2 vaccine as part of a routine program, were included in this study. The exclusion criteria were as follows: 1) cases that we could not obtained informed consent, 2) cases with past Covid-19 infection, 3) cases whose antibody titer before vaccination was elevated, 4) cases with new Covid-19 infection after vaccination, and 5) cases who could not provide a blood sample within the expected deadline (i.e., twenty days post-vaccination).

### Data sources and measurements

A 5-ml blood sample was drawn from the intermediate cubital vein, and we determined the anti-SARS-CoV-2 antibody titer (Elecsys^®^ Anti-SARS-CoV-2 S RUO, Roche Diagnostics K.K.) a) before vaccination (baseline), b) seven to twenty days after the first vaccination (first antibody test), and c) seven to twenty days after the second vaccination (second antibody test). We performed biochemical examinations (AST: aspartate aminotransferase, ALT: alanine aminotransferase, γ-GT: γ-glutamyl transpeptidase, Alb: albumin, TG: triglyceride, HDL-C: high density lipoprotein cholesterol, LDL-C: low density lipoprotein cholesterol, Cr: Creatinine, BS: Blood Sugar, HbA1c: Hemoglobin A1c, CRP: C - reactive protein) and we also calculated the blood cell count (White Blood Cells, Hemoglobin, Hematocrit, and Platelets) before vaccination. Self-reports were used to register participants’ information with respect to their age, gender, height, body weight, body mass index (BMI), past medical histories (1. Chronic lung disease (Chronic obstructive pulmonary disease: COPD), 2. Chronic lung disease (non-COPD), 3. Cardiac disease, 4. Hypertension, 5. Diabetes, 6. Dyslipidemia, 7. Liver disease, 8. Chronic kidney disease: CKD, 9. Autoimmune disease, 10. Cancer), medication (1. Antihypertensive drug, 2. Antidiabetic drug, 3. Antilipid drug, 4. Antiplatelet and Anticoagulant drug, 5. Immunosuppressive drug, 6. Immunoglobulin), smoking habits (current and past smoking history), habits of drinking alcohol, sleep duration per day, the quality of sleep (good or disturbed), number of meals per day, the quality of meal (good or disturbed), exercise hours per day, the number of outdoor activity days per week, and the length of days for vaccination and antibody test.

The diagnostic definition of the above diseases conformed to the WHO guideline, and we defined obesity as follows: 1) Underweight: BMI < 18.5, 2) Normal range: 18.5 ≦ BMI < 25, 3) Pre-obese: 25 ≦ BMI < 30, 4) Obese class I: 30 ≦ BMI < 35, 5) Obese class II: 35 ≦ BMI < 40, 6) Obese class III: 40 ≦ BMI (***World Health Organization, 2020***).

The low-antibody titer group (LABG) was defined as the group having less than 25 percentiles of antibody titer, whereas non-LABG was defined as having more than 25 percentiles of antibody titer.

### Statistical analysis

A sample size of 460 participants was determined based upon 70% power, 0.05 significance level, 0.3 effect size, three allocation ration, and 20% attrition. Unadjusted analysis evaluated between low and non-low-antibody titer group using the Student’s *t* test and Mann–Whitney *U* test for continuous variables, which were described as medians and interquartile ranges (IQR), and Fisher’s exact test or Pearson’s χ^2^ test for categorical variables, which were described as numbers and percentages. Univariate and Multivariate logistic regression analysis was performed to ascertain the effects of factors on the likelihood of LABG. Odds ratios and corresponding 95% confidence intervals were calculated. A *p* value of less than 0.05 was considered to indicate statistical significance. Data were analyzed with the Statistical Package for the Social Sciences, version 26.0 (SPSS, Chicago, IL, USA).

## Results

Of the 501 participants who fulfilled the inclusion criteria of this study, we excluded 96 participants because we did not obtain consents, seven participants because of past Covid-19 infection, one participant because of antibody titer elevation prior to the vaccination, one participant because of new Covid-19 infection after vaccination, and 22 participants because they could not provide a blood sample within the respective deadline. Finally, 374 participants were analyzed that were divided into 94 LABG and 280 non-LABG (Fig 1).

**Figure 1.**
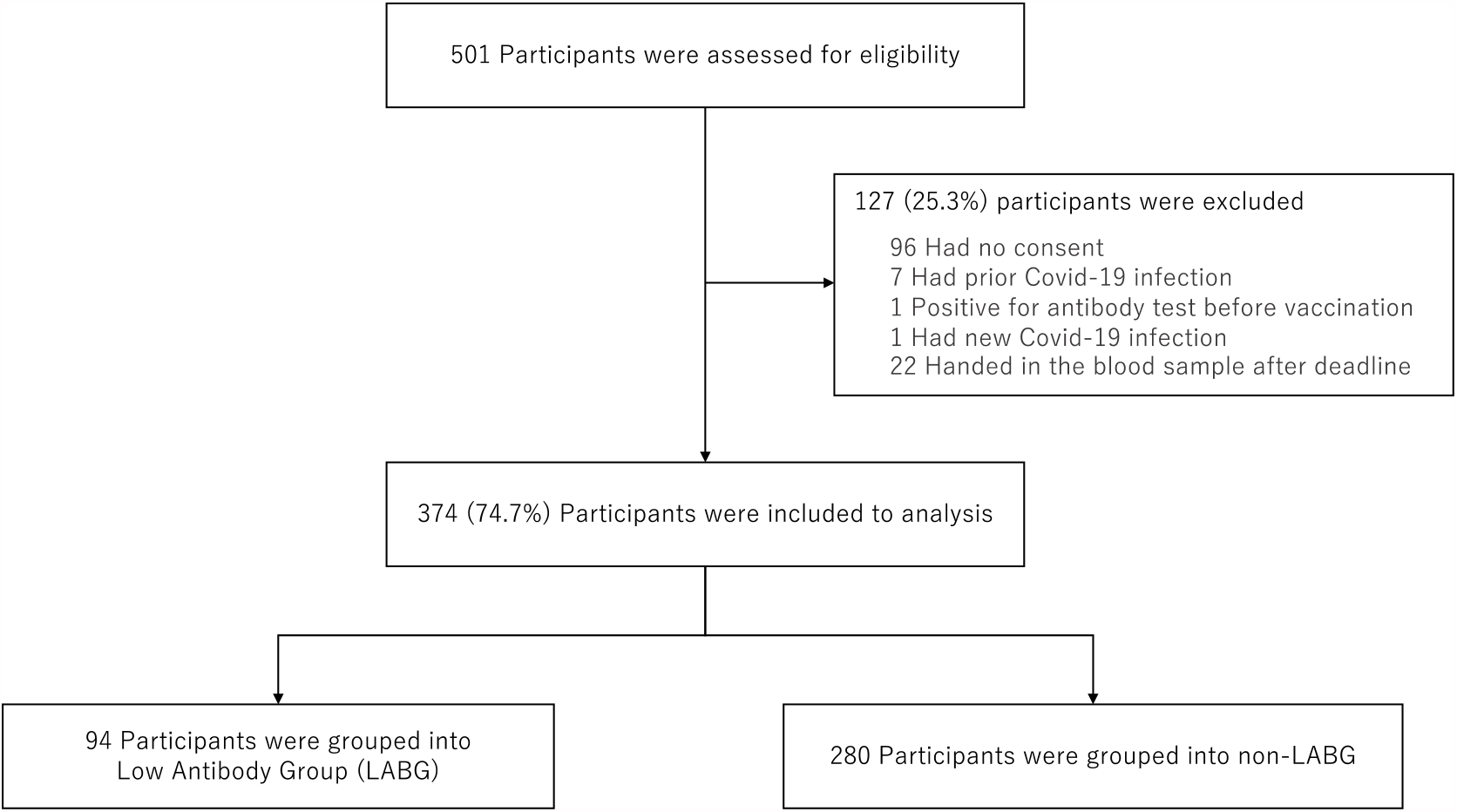
Flowchart for sample selection.

The baseline characteristics are shown in Table 1. The median age (interquartile range) of the participants was 36 (16.0) years, and 110 (29.4%) participants were males. Furthermore, 53 (14.2%) participants with pre-obesity (BMI: 25− 30) and 19 (5.0%) participants with obesity (BMI: >30) and zero participants belonging in the obese class III were identified. With respect to comorbidities, 118 (31.6%) participants had past medical histories, chronic lung disease (35 cases: 9.4%), hypertension (30 cases: 8.0%), and dyslipidemia (16 cases: 4.3%). The median duration (interquartile range) between the time from the first vaccination to the first antibody test, and from the second vaccination to the second antibody test was eight (1) days. Moreover, the median length of days from the first to the second vaccination was 22 (3) days.

**Table 1.** Baseline characteristics of the study population.

|  | Total Population (n=374)<br>Median (interquartile range) |
| --- | --- |
| Age, years | 36 (16.0) |
| Sex [n (%)] |  |
| Male | 110 (29.4) |
| Female | 264 (70.6) |
| Body Mass Index (BMI), kg/m<sup>2</sup> |  |
| BMI < 18.5 | 34 (9.1) |
| 18.5 ≤ BMI <25 | 268 (71.7) |
| 25 ≤ BMI <30 | 53 (14.2) |
| 30 ≤ BMI <35 | 14 (3.7) |
| 35 ≤ BMI <40 | 5 (1.3) |
| Past Medical Histories [n (%)] |  |
| Chronic lung disease (COPD) | 1 (0.3) |
| Chronic lung disease (non-COPD) | 34 (9.1) |
| Cardiac disease | 6 (1.6) |
| Hypertension | 30 (8.0) |
| Diabetes | 6 (1.6) |
| Dyslipidemia | 16 (4.3) |
| Liver disease | 6 (1.6) |
| Chronic kidney disease | 4 (1.1) |
| Autoimmune disease | 7 (1.9) |
| Cancer | 8 (2.1) |
| Medication [n (%)] |  |
| Antihypertensive drug | 28 (7.5) |
| Antidiabetic drug | 5 (1.3) |
| Antilipid drug | 13 (3.5) |
| Anticoagulant / Antiplatelet drug | 2 (0.5) |
| Immunosuppressive drug | 3 (0.8) |
| Immunoglobulin | 4 (1.1) |
| Current smoking [n (%)] | 70 (18.7) |
| Past smoking [n (%)] | 64 (17.1) |
| Drinking Alcohol [n (%)] | 229 (61.2) |
| Laboratory test |  |
| WBC | 6.35 (2.0) |
| Hb | 13.9 (1.7) |
| Hct | 40.2 (4.5) |
| PLT | 25.2 (7.3) |
| Alb | 4.6 (0.4) |
| AST | 19.0 (6.0) |
| ALT | 15.0 (12.0) |
| γ-GT | 17.0 (12.8) |
| Cr | 0.6 (0.2) |
| CRP | 0.03 (0.05) |
| BS | 90 (13.0) |
| HbA1c | 5.5 (0.4) |
| HDL-C | 65.0 (19.0) |
| LDL-C | 107 (35.0) |
| TG | 74.0 (59.0) |
| Sleep duration (SD) [n (%)] |  |
| SD ≤ 2 hours | 1 (0.3) |
| 3 hours ≤ SD < 4 hours | 64 (17.1) |
| 5 hours ≤ SD < 7 hours | 297 (79.4) |
| 8 hours ≤ SD < 10 hours | 12 (3.2) |
| The quality of sleep [n (%)] |  |
| Good | 232 (62.0) |
| Disturbed | 142 (38.0) |
| The number of meals per day [n (%)] |  |
| Once | 1 (0.3) |
| Twice | 104 (27.8) |
| More than 3 times | 269 (71.9) |
| The exercise hours (EH) per day [n (%)] |  |
| EH < 30 minutes | 271 (72.5) |
| 30 minutes ≤ EH < 1 hour | 79 (21.1) |
| 1 hour ≤ EH | 24 (6.4) |
| The numbers of outdoor activity days per week [n (%)] |  |
| No | 199 (53.2) |
| More than once | 175 (46.8) |
| The length of days for vaccination and antibody test, days |  |
| 1st vaccination to 1st antibody test | 8 (1) |
| 2nd vaccination to 2nd antibody test | 8 (1) |
| 1st vaccination to 2nd vaccination | 22 (3) |

Data are presented as the median (interquartile range) for continuous variables and the number (%) for categorical variables. COPD, Chronic obstructive pulmonary disease; WBC, White blood cells; Hb, Hemoglobin; Hct, Hematocrit; PLT, Platelets; Alb, Albumin; AST, Aspartate aminotransferase; ALT, alanine aminotransferase; γ-GT, γ-glutamyl transpeptidase; Cr, Creatinine; CRP, C-reactive protein; BS, Blood sugar; HbA1c, Hemoglobin A1c; HDL-C, High density lipoprotein cholesterol; LDL-C, Low density lipoprotein cholesterol; TG, Triglyceride.

Almost all antibody titer was not elevated in the first antibody test, and only 23 (6.2%) samples exhibited a slight positive antibody response, with a mean value of 0.41 U/mL. All samples showed significant antibody elevation in the second antibody test, with a mean value of 3476 U/mL (Fig 2).

**Figure 2.**
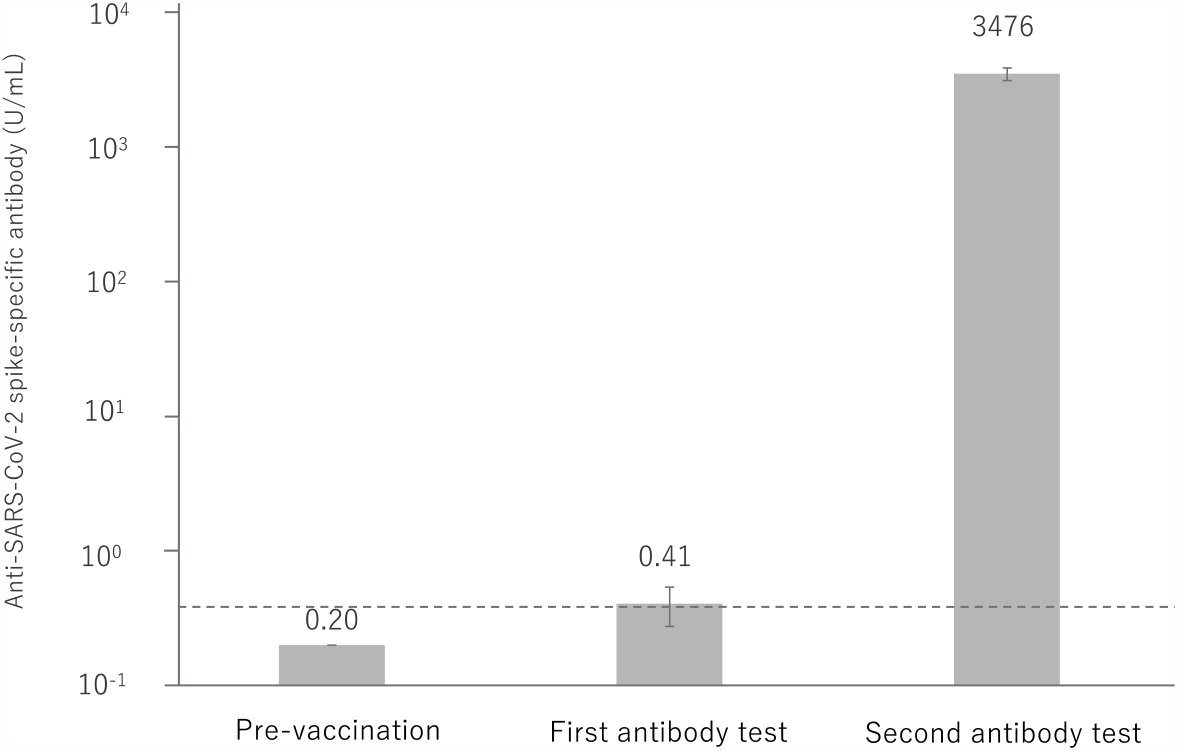
Anti-SARS-CoV-2 spike-specific antibody response to BNT162b2 vaccination. 374 participants were administered the BNT162b2 vaccine. Serum samples were obtained before injection and 7 to 20 days after the first and second vaccination. Each bar shows the geometric mean concentrations of anti-SARS-CoV-2 spike-specific antibody (lower limit of quantitation, 0.40; dashed line). The top of the vertical bar represents the mean with a 95% confidence interval (I bar). For values below the lower limit of quantification (LLOQ)=0.40, LLOQ/2 were included in the calculation.

Table 2 shows the comparison between LABG and non-LABG. The median age and HbA1c were significantly higher in the LABG group (p < 0.001, < 0.01). The proportion of males (62 (33.2%) vs females (48 (25.7%)) was greater in the LABG group, but with no significant differences (p=0.38). Moreover, the proportion of participants with low BMI (<18.5) and high BMI (>30) were significantly higher in the LABG group (p < 0.05). The proportion of chronic lung disease, hypertension, diabetes, dyslipidemia, autoimmune disease, and cancer was significantly higher in the LABG group (p < 0.05). Although no significant difference was confirmed in the hours of exercise per day, the proportion of participants with no-outdoor activities was significantly higher in the LABG group (p < 0.001). Finally, the length of days from the second vaccination to the second antibody test, and from the first to the second vaccination was significantly longer in the non-LABG group (p < 0.05, < 0.05).

**Table 2.**
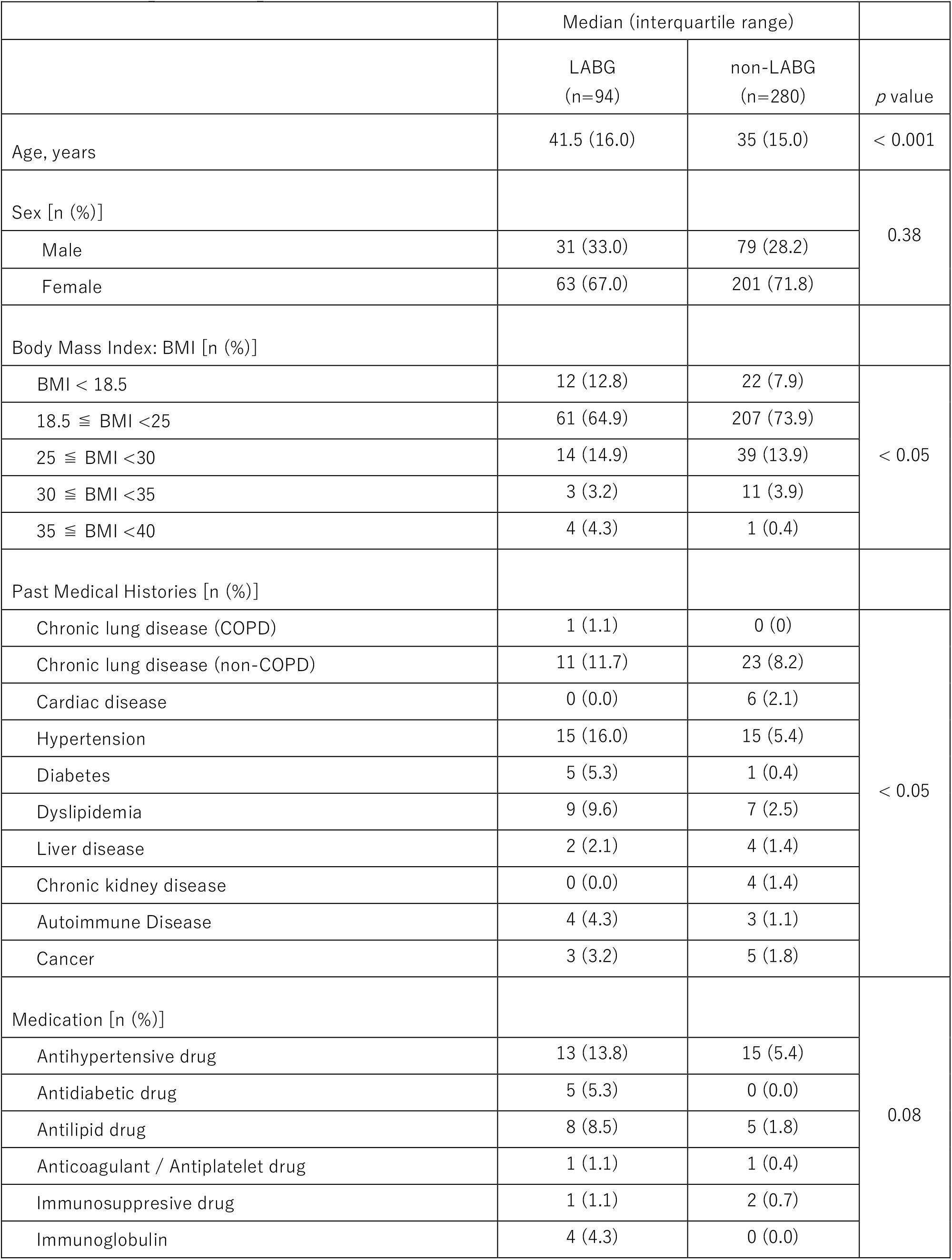
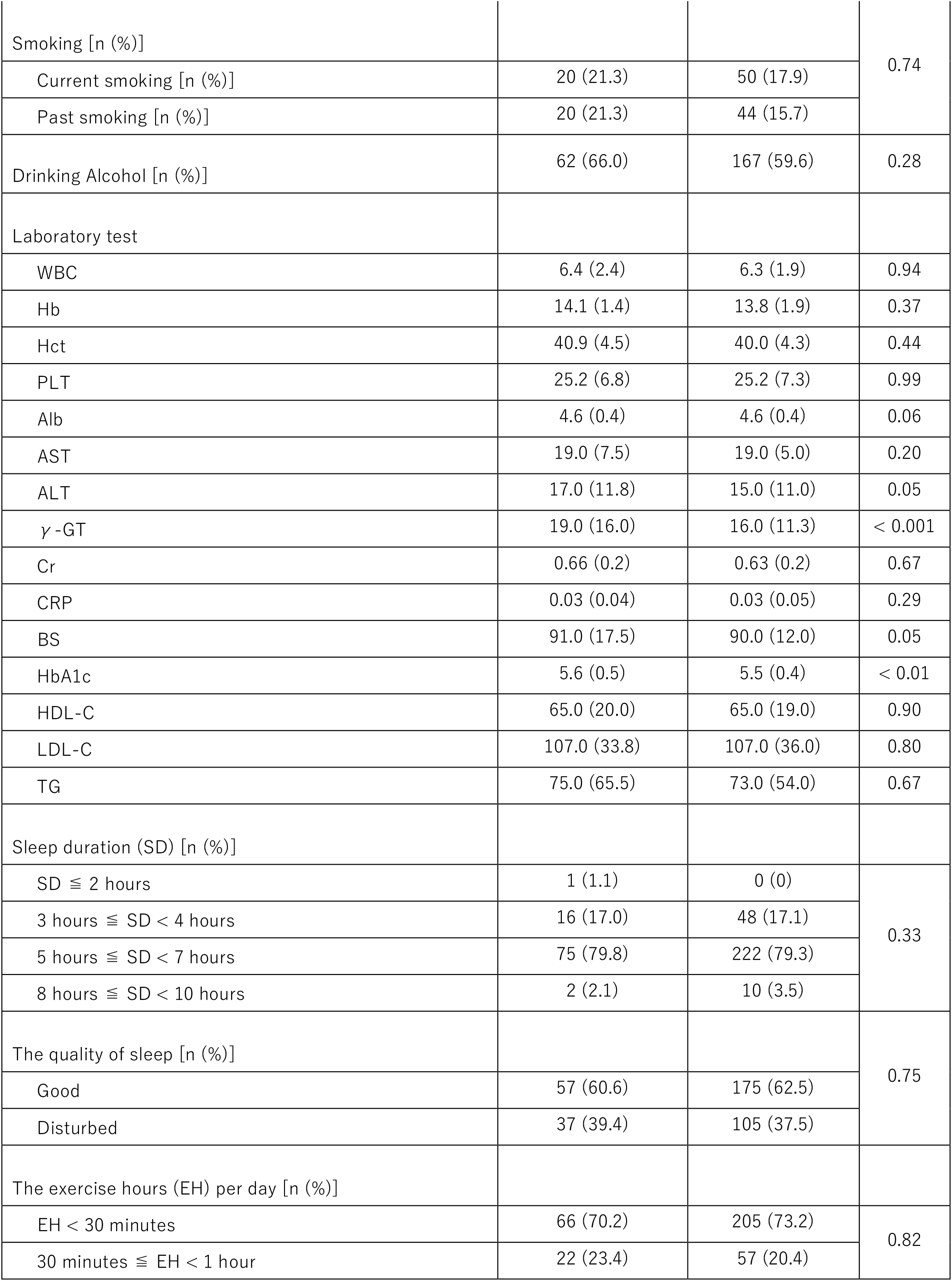
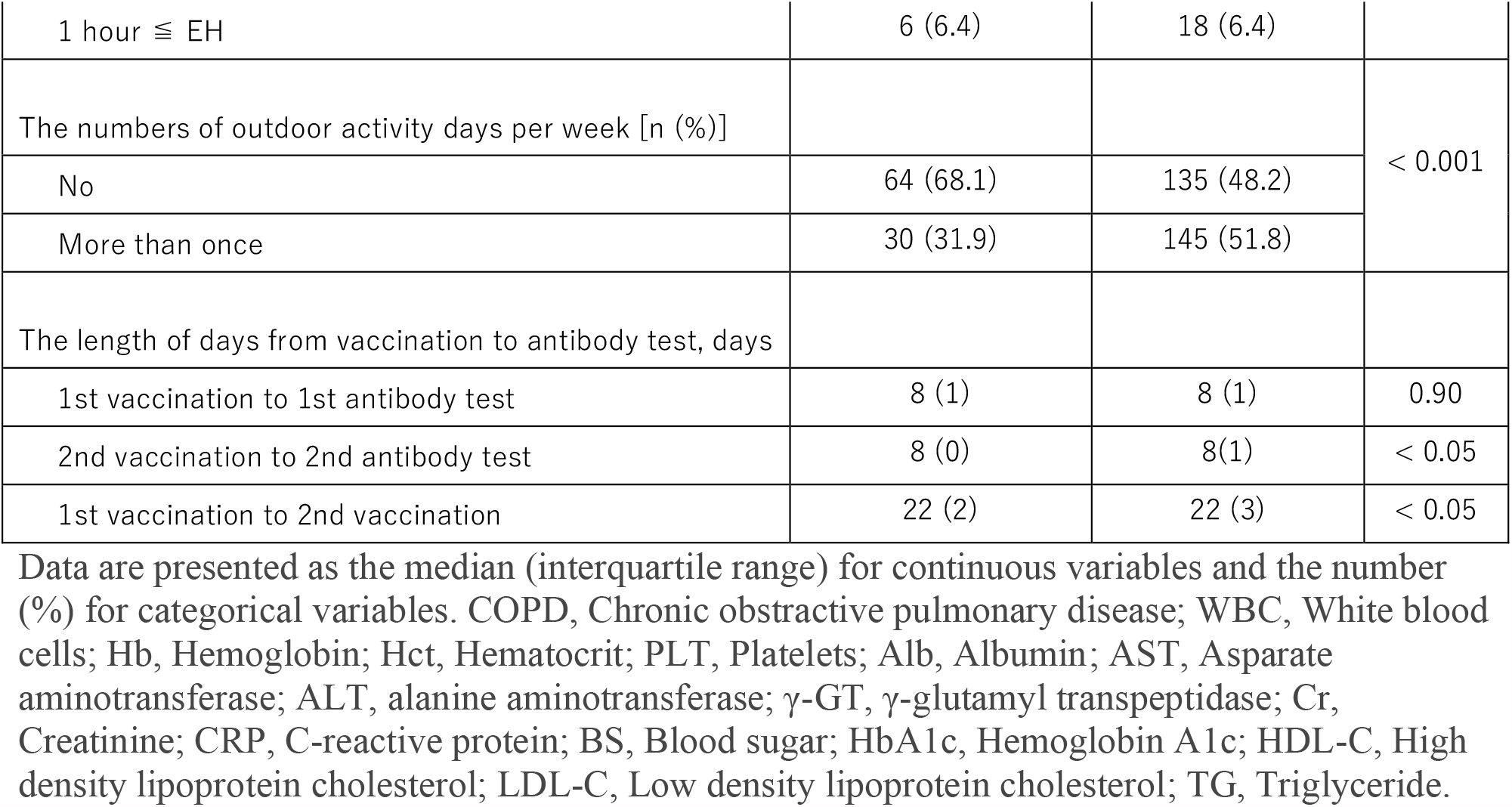
Comparison of parameters between LABG and non-LABG.

Table 3 shows the univariate and multivariate logistic regression analysis of the factors associated with LABG. Age, obesity (obese class II), hypertension, diabetes, dyslipidemia, antihypertensive drug, antilipid drug, γ-GT, BS, HbA1c, and lack of outdoor activity were a significant suppressor of the antibody response. The univariate logistic regression analysis revealed that the length of days from the first to the second vaccination promoted a significant antibody response.

**Table 3.**
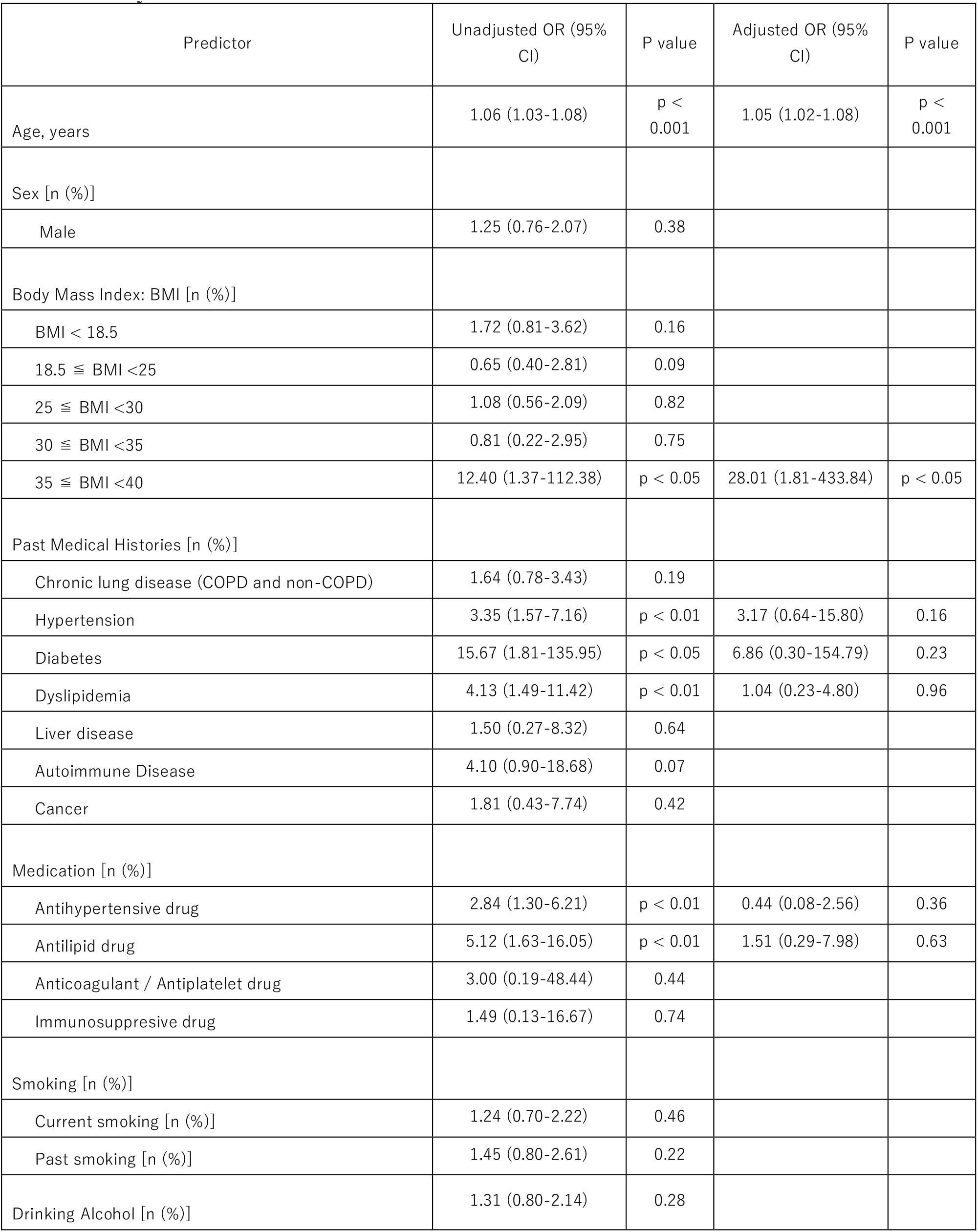
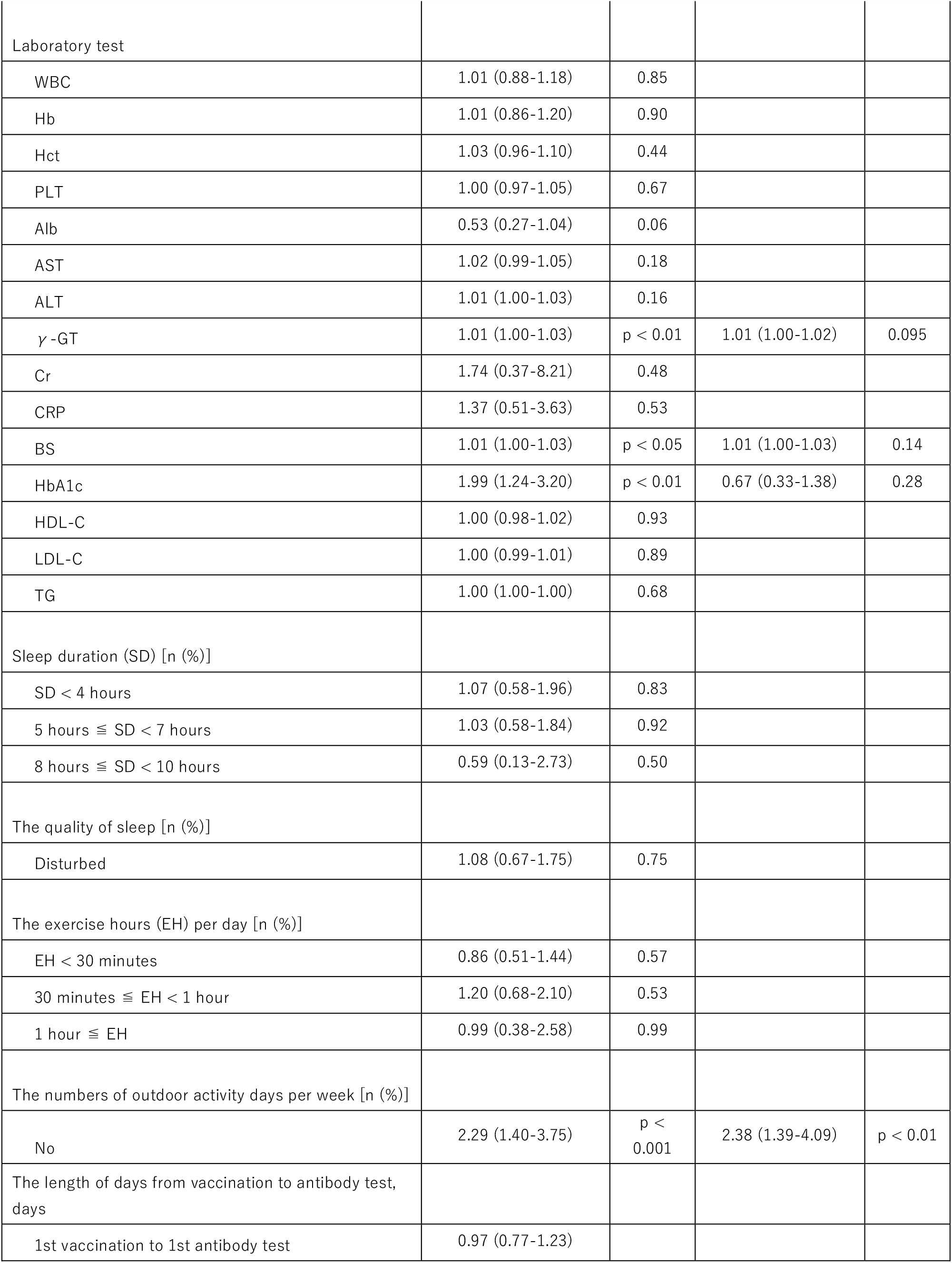
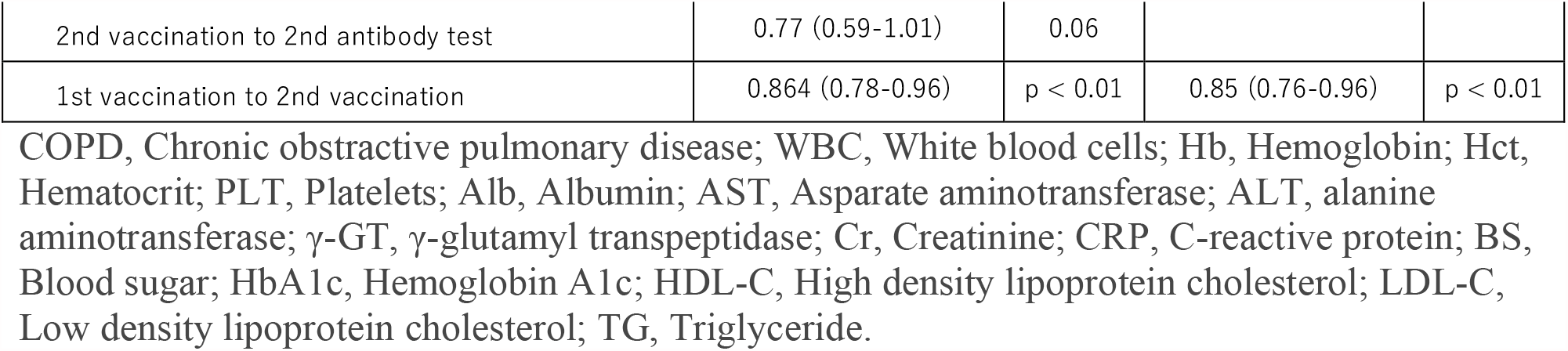
Univariate and multivariate logistic regression analysis of factors associated with low-antibody titer after vaccination.

| Predictor | Unadjusted OR (95% CI) | P value | Adjusted OR (95% CI) | P value |
| --- | --- | --- | --- | --- |
| Age, years | 1.06 (1.03-1.08) | p < 0.001 | 1.05 (1.02-1.08) | p < 0.001 |
| Sex [n (%)] |  |  |  |  |
| Male | 1.25 (0.76-2.07) | 0.38 |  |  |
| Body Mass Index: BMI [n (%)] |  |  |  |  |
| BMI < 18.5 | 1.72 (0.81-3.62) | 0.16 |  |  |
| 18.5 ≤ BMI < 25 | 0.65 (0.40-2.81) | 0.09 |  |  |
| 25 ≤ BMI < 30 | 1.08 (0.56-2.09) | 0.82 |  |  |
| 30 ≤ BMI < 35 | 0.81 (0.22-2.95) | 0.75 |  |  |
| 35 ≤ BMI < 40 | 12.40 (1.37-112.38) | p < 0.05 | 28.01 (1.81-433.84) | p < 0.05 |
| Past Medical Histories [n (%)] |  |  |  |  |
| Chronic lung disease (COPD and non-COPD) | 1.64 (0.78-3.43) | 0.19 |  |  |
| Hypertension | 3.35 (1.57-7.16) | p < 0.01 | 3.17 (0.64-15.80) | 0.16 |
| Diabetes | 15.67 (1.81-135.95) | p < 0.05 | 6.86 (0.30-154.79) | 0.23 |
| Dyslipidemia | 4.13 (1.49-11.42) | p < 0.01 | 1.04 (0.23-4.80) | 0.96 |
| Liver disease | 1.50 (0.27-8.32) | 0.64 |  |  |
| Autoimmune Disease | 4.10 (0.90-18.68) | 0.07 |  |  |
| Cancer | 1.81 (0.43-7.74) | 0.42 |  |  |
| Medication [n (%)] |  |  |  |  |
| Antihypertensive drug | 2.84 (1.30-6.21) | p < 0.01 | 0.44 (0.08-2.56) | 0.36 |
| Antilipid drug | 5.12 (1.63-16.05) | p < 0.01 | 1.51 (0.29-7.98) | 0.63 |
| Anticoagulant / Antiplatelet drug | 3.00 (0.19-48.44) | 0.44 |  |  |
| Immunosuppressive drug | 1.49 (0.13-16.67) | 0.74 |  |  |
| Smoking [n (%)] |  |  |  |  |
| Current smoking [n (%)] | 1.24 (0.70-2.22) | 0.46 |  |  |
| Past smoking [n (%)] | 1.45 (0.80-2.61) | 0.22 |  |  |
| Drinking Alcohol [n (%)] | 1.31 (0.80-2.14) | 0.28 |  |  |
| Laboratory test |  |  |  |  |
| WBC | 1.01 (0.88-1.18) | 0.85 |  |  |
| Hb | 1.01 (0.86-1.20) | 0.90 |  |  |
| Hct | 1.03 (0.96-1.10) | 0.44 |  |  |
| PLT | 1.00 (0.97-1.05) | 0.67 |  |  |
| Alb | 0.53 (0.27-1.04) | 0.06 |  |  |
| AST | 1.02 (0.99-1.05) | 0.18 |  |  |
| ALT | 1.01 (1.00-1.03) | 0.16 |  |  |
| γ-GT | 1.01 (1.00-1.03) | $p < 0.01$ | 1.01 (1.00-1.02) | 0.095 |
| Cr | 1.74 (0.37-8.21) | 0.48 |  |  |
| CRP | 1.37 (0.51-3.63) | 0.53 |  |  |
| BS | 1.01 (1.00-1.03) | $p < 0.05$ | 1.01 (1.00-1.03) | 0.14 |
| HbA1c | 1.99 (1.24-3.20) | $p < 0.01$ | 0.67 (0.33-1.38) | 0.28 |
| HDL-C | 1.00 (0.98-1.02) | 0.93 |  |  |
| LDL-C | 1.00 (0.99-1.01) | 0.89 |  |  |
| TG | 1.00 (1.00-1.00) | 0.68 |  |  |
| Sleep duration (SD) [n (%)] |  |  |  |  |
| SD < 4 hours | 1.07 (0.58-1.96) | 0.83 |  |  |
| 5 hours ≤ SD < 7 hours | 1.03 (0.58-1.84) | 0.92 | | |
| 8 hours ≤ SD < 10 hours | 0.59 (0.13-2.73) | 0.50 | | |
| The quality of sleep [n (%)] |  |  |  |  |
| Disturbed | 1.08 (0.67-1.75) | 0.75 |  |  |
| The exercise hours (EH) per day [n (%)] |  |  |  |  |
| EH < 30 minutes | 0.86 (0.51-1.44) | 0.57 |  |  |
| 30 minutes ≤ EH < 1 hour | 1.20 (0.68-2.10) | 0.53 | | |
| 1 hour ≤ EH | 0.99 (0.38-2.58) | 0.99 | | |
| The numbers of outdoor activity days per week [n (%)] |  |  |  |  |
| No | 2.29 (1.40-3.75) | $p < 0.001$ | 2.38 (1.39-4.09) | $p < 0.01$ |
| The length of days from vaccination to antibody test, days |  |  |  |  |
| 1st vaccination to 1st antibody test | 0.97 (0.77-1.23) |  |  |  |
| 2nd vaccination to 2nd antibody test | 0.77 (0.59-1.01) | 0.06 |  |  |
| 1st vaccination to 2nd vaccination | 0.864 (0.78-0.96) | p < 0.01 | 0.85 (0.76-0.96) | p < 0.01 |

COPD, Chronic obstructive pulmonary disease; WBC, White blood cells; Hb, Hemoglobin; Hct, Hematocrit; PLT, Platelets; Alb, Albumin; AST, Asparate aminotransferase; ALT, alanine aminotransferase; γ-GT, γ-glutamyl transpeptidase; Cr, Creatinine; CRP, C-reactive protein; BS, Blood sugar; HbA1c, Hemoglobin A1c; HDL-C, High density lipoprotein cholesterol; LDL-C, Low density lipoprotein cholesterol; TG, Triglyceride.

In addition, the multivariate logistic regression analysis, showed that age, obesity (obese class II), and lack of outdoor activity were significant suppressors of the antibody response, whereas the length of days from the first to the second vaccination was promoted a significant antibody reaction.

## Discussion

Our data showed that the antibody titer was dramatically elevated after the second administration of the BNT162b2 vaccine in Japanese people. Moreover, our study identified that older age, obesity (obese class II), and performing no-outdoor activities were significant suppressors of antibody response, as opposed to the duration between the first and the second vaccination which were found to promote a significant antibody response. This is the first study that evaluated the antibody response in a relatively large cohort following mRNA vaccination for SARS-CoV-2 in Japan, and assessed various factors in terms of promoting or suppressing a subsequent antibody response.

According to previous studies, the median antibody titers were 9,136 or 27,872 U/ml at seven days post-administration of the second vaccination, and 0.6 or 0.8 U/ml at seven days post-administration of the first vaccination (***Walsh et al., 2020; Mulligan et al., 2020***). In the present study, participants demonstrated a 6.2% and 100% positive antibody reaction for SARS-CoV-2 in the first and the second antibody test, respectively. The median antibody titers were 1.5 U/mL in the first antibody test and 2620 U/mL in the second antibody test, values which are consistent with current literature. Our findings demonstrate that the second dose of the BNT162b2 two-dose vaccine can efficiently enhance the immunity of Asian people.

Several studies and guidelines showed that age, male, obesity, diabetes, chronic lung disease, hypertension, dyslipidemia, chronic kidney disease and smoking were risk factors for the development of severe Covid-19 (***Matsunaga et al., 2020; Mi et al., 2020; Liang et al., 2020; Lippi et al., 2020; Myers et al., 2020; Fadini et al., 2020; Zheng et al., 2020; Popkin BM et al., 2020***). A study performed by Pellini *et al* demonstrated that obesity, age and male sex may be hampering SARS-CoV-2 vaccine immunogenicity (***Pellini et al., 2021***), but there was no other similar study for comparison between vaccine immunogenicity and the factors affecting antibody response.

Obesity, which also causes diabetes and hypertension, is one of the most important risk factors of severe Covid-19 by suppressing immune system responses. In this study, we defined obesity in accordance with the WHO guidelines (***World Health Organization, 2020***). A common definition of obesity is important for comparing results of studies performed in different countries, especially when it comes to the identification of immune responses pertaining to racial differences. A previous review by Millner *et al* showed that the excess adipose tissue could block the supply of nutrients to immune cells (***Milner et al., 2012***). Moreover, a study performed by Vandanmagsar *et al* revealed that low levels of inflammation and relatively high levels of inflammatory cytokines induced by adipose tissue can decrease immune responses, and especially T lymphocyte activity (***Vandanmagsar et al., 2011***). These immune cell suppression mechanisms can reduce the subsequent antibody production. Similar to these previous studies, our data demonstrated that the rates of obesity (obese class II) were significantly larger in the LABG group. Compared to the group with BMI of 35 or less, obesity (obese class II) had a significantly higher odds ratio for low-antibody response, with a value of 28.01. Another important question pertaining to obese participants was whether the vaccine solution was actually injected into the muscle tissue. In the muscle tissue, the blood stream is abundant, large numbers of immune cells are distributed in the muscle, thus the subsequent activation of immunity by the vaccine can occur more easily. Moreover, intramuscular injection, as opposed to subcutaneous injection, is definitely more critical for mRNA vaccines, because mRNA vaccines are so fragile that they need to be quickly recognized by immune cells (***Zeng et al., 2020***). According to the research by Akkus *et al* (***Akkus et al., 2012***), the maximum subcutaneous adipose tissue thicknesses was approximately 18 mm for triceps, even when participants’ BMI is more than 30 kg/m^2^. Furthermore, Takahashi *et al* presented a calculation formula for the subcutaneous tissue thickness for triceps (Y) as follows; 1) male: Y(cm) = 0.04X− 0.25 and 2) female: Y(cm) = 0.04X− 0.17 (X = BMI) (***Takahashi et al., 2014***). The length of the needle we used for vaccination was 25 mm, hence it was very unlikely that the needle would not reach the muscle.

Similar to obesity, diabetes is also a significant risk factor element for the development of severe Covid-19, and a report by Fadini *et al* showed that the presence of diabetes increases the risk by 2.3 times (***Fadini et al., 2020***). In addition, Berbudi *et al* showed that hyperglycemia could inhibit IL-6 production, which induces antibodies and T cells (***Berbudi et al., 2020***), and that this was a mechanism that triggered low-antibody production, similar to obesity. Our study showed that the rates of diabetes, median blood sugar, and median HbA1c were significantly higher in the LABG group. However, the subsequent adjustment did not reveal any significant differences between the two groups.

Gustafson *et al* exhibited that the immune response toward vaccination is controlled by a delicate balance of effector T cells and follicular T cells, yet the aging process disturbs this balance.

Several changes in T cells have been identified that contribute to age-related defects of post-transcriptional regulation, T cell receptor signaling, and metabolic function (***Gustafson et al., 2020***). Similar to Pellini’s study, our study revealed that the antibody titer after mRNA SARS-CoV-2 vaccine significantly reduces as older.

The Japanese Covid-19 guideline does not include gender in the risk factors for severe Covid-19, but a report published overseas indicated that male patients are approximately three times more likely to be admitted to intensive care units and have a higher mortality rate (***Mi et al., 2020***).

Females are known to be sensitive to immune responses, including antibody production to infectious diseases, which in turn induces the development of autoimmune diseases (***Fischinger et al., 2019***).

Differences in sex hormones are associated with gender differences in vaccine-induced immunity. For example, testosterone levels and Influenza vaccine antibody titer have been shown to be inversely correlated (***Markle et al., 2014; Ruggieri et al., 2016; Furman et al., 2014***). Genetic differences, as well as sex hormone differences, affect vaccine-induced immunity. The X chromosome expresses 10 times more genes than the Y chromosome, and differences in gene expression between the X and Y chromosomes promote differences in vaccine-induced immunity by gender (***Fischinger S et al., 2019***). Our study showed that although not significant the proportion of males was larger in the LABG group, a finding that agrees with current literature.

In contrast, several studies highlighting the relationship between smoking and vaccine-induced immunological reactions, have been reported. However, the results of these studies vary and none of them explains the underlying mechanism with which smoking influences antibody production (***Winter et al., 1994; Namujju et al., 2014***). Certain studies showed that smoking induced inflammatory cytokines and chemokines, but the effect of the complex mixture of chemicals in tobacco varies depending on the individual smoking habits (***Kubo et al., 2005; Daloee et al., 2017***). In our study, we did not identify any significant difference in smoking between the LABG and non-LABG groups.

Furthermore, and although there was no difference in the exercise hours between the two groups, including indoor activities, our study revealed that the proportion of no-outdoor activity days was significantly larger in the LABG group. Moreover, participants that did not perform outdoor activities had a significantly higher odds ratio, which was 2.38 after adjustment for LABG. Researchers have long focused on the effect of vitamin D on the activation of the immune system, and a study carried by Kashi *et al* showed that there was a positive correlation between plasma vitamin D levels and post-vaccination antibody titer for Hepatitis B (***Kashi et al., 2021***). Although no studies have further clarified the relationship between antibody titer and vitamin D levels after vaccination for Covid-19, Merzon *et al* found that low plasma vitamin D levels or short duration of exposure to sunlight were associated with an increased risk of Covid-19 infection (***Merzon et al., 2020***). Therefore, vitamin D levels may be positively correlated with immune responses following administration of the BNT162b2 vaccine.

Two major studies showed that sleep duration and sleep quality were positively correlated to antibody production after Influenza or Hepatitis A vaccination, respectively (***Tanja et al., 2003; Prather et al., 2021***). However, to our knowledge, there have been no other studies that have evaluated the effect of sleep to the triggering of antibody reactions after Covid-19 vaccination. In our study, we could not show any significant relationship between sleep and antibody response. Parry et al. showed that the antibody response was 3.5-fold higher in cases of delayed second vaccination dose (12 weeks after he first vaccination), and the cellular immune responses were 3.6-fold lower compared to cases with normal vaccination schedule (***Parry et al., 2021***). Our study revealed that a longer interval between the first and the second vaccination had a significant positive correlation with antibody response.

The antibody titer level does not necessarily reflect the immune function against pathogens. However, our study showed that lifestyle improvements such as performing outdoor activities and reducing complications such as obesity may increase the immune response of the BNT162b2 vaccine, and this fact is especially important for people with risk factors for severe Covid-19. In addition to this, further research of the vaccination schedule is also needed for promoting immune responses after BNT162b2 vaccine administration, especially to older people.

Our study has several limitations. First, we only diagnosed new Covid-19 infections to participants through symptoms of fever and common cold. Therefore, we were unable to identify asymptomatic infections. However, our data showed that the number of antibody-positive cases at baseline, without any previous episode of Covid-19 infection was only one (0.3%), and thus had low statistical power. Second, we only calculated the antibody titer but not the CD4 lymphocyte activity. Consequently, we could not evaluate the effect of these factors to the entire immune system. Third, we only included relatively healthy medical workers, but did not include participants with severe complications, and this may have reduced the difference between the LABG and non-LABG groups. Further large-scale studies that include participants with several concurrent conditions are needed in the future.

## Conclusions

Our single-center study demonstrated that age, obesity (obese class II), and lack of outdoor activities were significant suppressors of antibody responses, whereas the length of days from the first to the second vaccination promoted a significant antibody response. Evidence from multi-center studies is needed to develop further vaccination strategies.

## Supporting information

https://www.amazon.co.jp/clouddrive/share/5bIyDUe1rQDceyLqMaKp5Jg4C70DOeC05y760tZvYRE

## Data Availability

We accepted to show all the data set.

https://www.amazon.co.jp/clouddrive/share/5bIyDUe1rQDceyLqMaKp5Jg4C70DOeC05y760tZvYRE

## Acknowledgements

None

